# Modeling the evolution of SARS-CoV-2 under non-pharmaceutical interventions

**DOI:** 10.1101/2021.02.20.21252133

**Authors:** Yael Gurevich, Yoav Ram, Lilach Hadany

## Abstract

Social and behavioral non-pharmaceutical interventions (NPIs), such as mask-wearing, social distancing, and travel restrictions, as well as diagnostic tests, have been broadly implemented in response to the COVID-19 pandemic. Epidemiological models and data analysis affirm that wide adoption of NPIs helps to control the pandemic. However, SARS-CoV-2 has extensively demonstrated its ability to evolve. Therefore, it is crucial to examine how NPIs may affect the evolution of the virus. Such evolution could have important effects on the spread and impact of the pandemic.

We used evo-epidemiological models to examine the effect of non-pharmaceutical interventions and testing on two evolutionary trajectories for SARS-CoV-2: attenuation and test evasion. Our results show that when stronger measures are taken, selection may act to reduce virulence. Additionally, the timely application of NPIs could significantly affect the competition between viral strains, favoring reduced virulence. Furthermore, a higher testing rate can select for a test-evasive viral strain, even if that strain is less infectious than the detectable competing strain. Importantly, if a less detectable strain evolves, epidemiological metrics such as confirmed daily cases may distort our assessment of the pandemic. Our results highlight the important implications NPIs can have on the evolution of SARS-CoV-2.

## Introduction

Social and behavioral non-pharmaceutical interventions (NPIs) have been broadly applied to contain the COVID-19 pandemic. These interventions include use of face masks, implementation of social distancing, closure of educational institutions, individual movement restrictions, and quarantining cases confirmed using RT-PCR or serological testing. The role of NPIs in controlling the COVID-19 pandemic has been studied extensively^1^. Epidemiological models^2–7^ have been used to assess the impact of these NPIs on the pandemic, aiming to forecast the levels of infection, hospitalization, and mortaility^8,9^. Both theoretical models^6,7^ and data analysis^10–13^ affirm that wide and early adoption of interventions, such as limiting social contacts and wearing face masks, helps to control the pandemic. However, SARS-CoV-2 has broadly demonstrated its ability to evolve^14^: it has been suggested that a mutation conferring ability to infect humans^15^ preceded its transmission to humans from bats^16^. Similar to other RNA viruses^17^, the mutation rate of SARS-CoV-2 is estimated at ∼10^−6^ per site/cycle, relatively high^18^ (although lower than influenza^19^). Additionally, there is already significant variation in the viral population^14,20^ due to a high rate of recombination^16^, a very high number of copies produced in each infection^18^, a rapid replication cycle (around 10 hours^18^), and the large effective size of the SARS-CoV-2 population. This variation can potentially lead to adaptive evolution^21^, as seen before, for example, in Influenza^22^, HIV^23^ and Ebola^24^

Because the virus only recently emerged in humans, further adaptation of SARS-CoV-2 to its new host is likely. Indeed, new strains have recently emerged carrying mutations that may increase transmission, lower detectability, and perhaps even reduce vaccine efficiency^25–27^. Importantly, by limiting the transmission of the virus, NPIs may exert strong selection on SARS-CoV-2^28^. Hence, it is crucial to examine how NPIs could affect the evolution of the virus. Such evolution may have important effects on the spread and control of the pandemic. To examine how the virus may evolve in response to NPIs, we have developed evo-epidemiological models^29,30^ that track both the infection status of the human hosts and the strain of the infecting virus. We use these models to examine how NPIs are expected to affect two evolutionary trajectories for SARS-CoV-2: attenuation and test evasion.

### Attenuation

An important epidemiological feature of the virus is the high frequency of asymptomatic infections^2,3,31–33^. It is suggested that asymptomatic individuals are infectious^34^ and can transmit the disease but are less infectious than those who are symptomatic^33,35^. Due to limited resources, the COVID-19 testing policy in many countries does not include routine screening of asymptomatic individuals, unless they have been in direct contact with a confirmed case or are routinely exposed to infected individuals (e.g., health workers). Thus, these asymptomatic cases largely go undetected. Asymptomatic infection allows the individual to maintain their normal routine and social contact levels throughout the entire course of the infection, thus potentially producing many secondary infections. The tendency to develop asymptomatic infection is affected both by the individual characteristics, such as prior health status^36^, and by the virus itself. As asymptomatic cases are less likely to be diagnosed and isolated, we hypothesized that a decrease in the frequency of symptomatic cases can be favored by selection, leading to the evolution of an attenuated pathogen^37^. For example, a mutation causing decreased viral load may cause a higher frequency of asymptomatic cases and milder disease. However, asymptomatic individuals are likely less infectious^33^, and if the relative transmissibility of asymptomatic cases is low enough, the more virulent strain may evolve. Increased awareness to the epidemic and application of NPIs may select for further increase in the frequency of asymptomatic infections. NPIs change the overall infection rate by reducing the number of contacts per individual, hence we expect that NPIs will have an important role in determining the outcome of competitions between attenuated and virulent strains.

### Test evasion

An active COVID-19 infection can be diagnosed using an RT-PCR test on a nasopharyngeal swab specimen^38^, detecting specific sites in the viral genome. The detected sites were chosen such that they are critical for virus function^39,40^. COVID-19 tests have been evaluated for their sensitivity, the expected fraction of infected individuals who receive a positive test result, and specificity, the fraction of uninfected individuals who erroneously receive a positive test result^41^. The conditions under which individuals are tested may differ between and even within different countries^42^. Given that individuals who receive a positive test result are isolated until recovery, largescale testing can exert strong selection pressure on the virus. The detectability of the virus may be directly under selection, potentially favoring two classes of mutants: (i) mutants presenting atypical infections, including affecting different age groups (e.g. children) or different tissues (e.g. gastrointestinal system^43^, heart and liver infections^44,45^). Undiagnosed COVID-19 patients may not be quarantined even when sick – heart disease, for example, is not usually infectious – and therefore might infect others, including healthcare workers and other patients. (ii) As PCR-based tests are used to determine who must be quarantined, selection might favor viruses with modifications in the RNA sequence used for the test^46^. We hypothesize that when testing is frequent and NPIs are significant, selection could favor strains that are harder to detect, even at the cost of lower transmissibility.

## Model

We use a SEIR compartmental epidemic model^2,3,10^. The model follows two viral strains simultaneously spreading in an initially susceptible population (Figure 1). We neglect births and deaths due to non-disease related causes and assume no superinfection and total cross-immunity, such that recovered individuals from either strain are immune to both strains^47^. For COVID-19, the latter is likely true in some strains for at least several months, due to the presence of antibodies^47^. Thus, we divide the host population to susceptible individuals (*S*), individuals exposed to one of the strains (*E*_1_, *E*_2_ for strain 1 and strain 2, respectively), asymptomatic 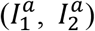, presymptomatic 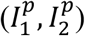, and symptomatic individuals 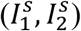, and “removed” individuals (*R*_1_, *R*_2_), which effectively include both recovered individuals and fatalities.

**Figure 1.**
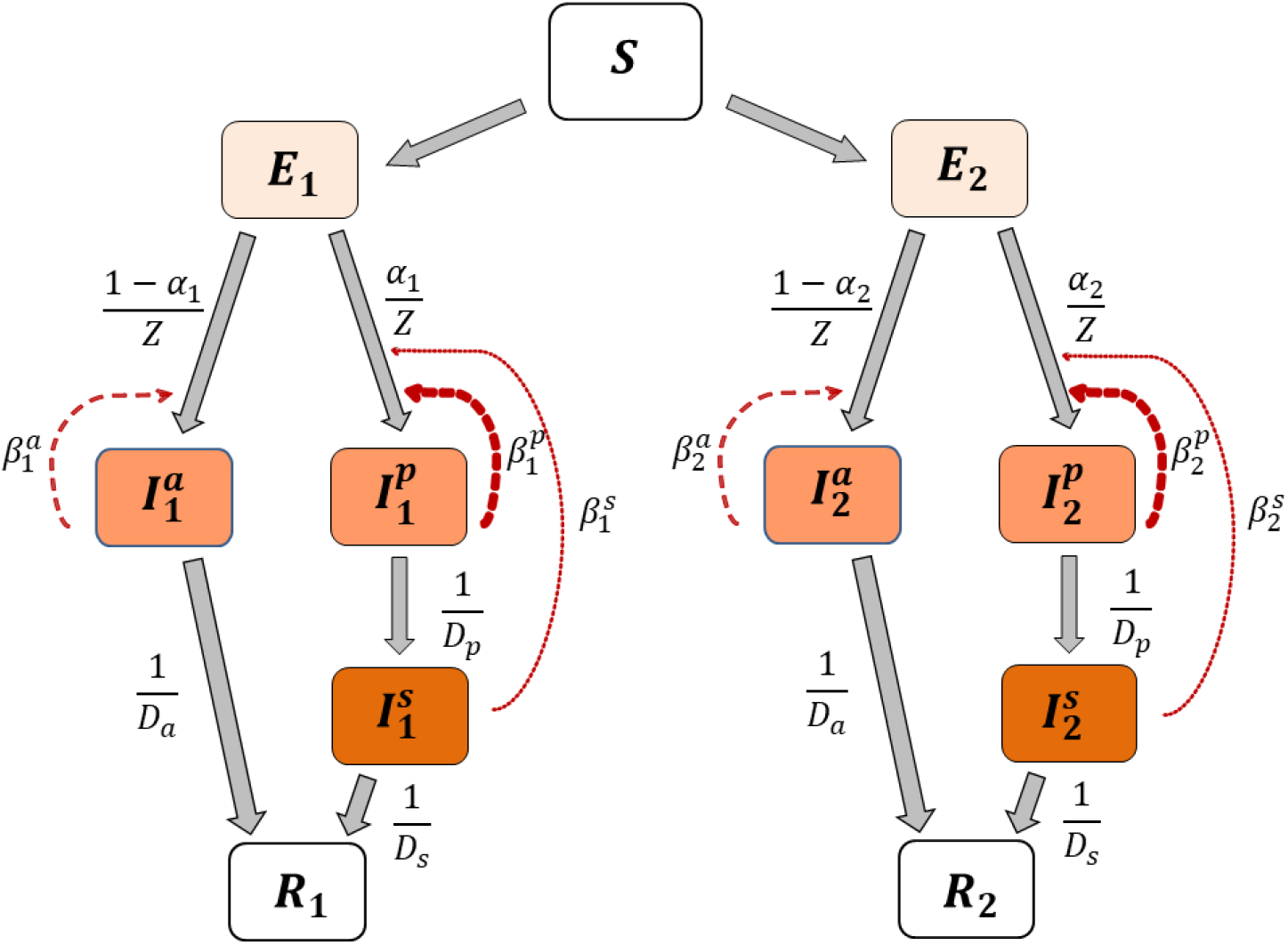
Evo-epidemiological model. Our model follows two viral strains simultaneously spreading in an initially susceptible population. Susceptible (*S*) individuals become exposed (*E*) after contact with an infected individual with rate *β*^*x*^ (where x=*p, s*, or *a* is the class of infected individual). Exposed individuals (*E*) undergo an incubation period during which they are not infectious. After an average incubation period of *Z* days, exposed individuals become infected, with either a presymptomatic (*I* ^*p*^) or asymptomatic (*I* ^*a*^) infection, with probability *α* and (1− *α*, respectively. Asymptomatic individuals (*I* ^*a*^) are infectious but asymptomatic from disease onset until recovery. Presymptomatic individuals (*I*^*p*^) are infectious and may be asymptomatic or exhibit mild symptoms for several days, after which they exhibit clinical manifestation of the disease (*I* ^*s*^) and are therefore isolated. Symptomatic and asymptomatic cases become recovered (*R*) after an average of *D*_*s*_ and *D*_*a*_ days, respectively.

Susceptible individuals become exposed through contact with an infected individual with rate *β*^*x*^ (where *x=p, s* or *a* is the class of infected individual). The distinction between *β*^*p*^,*β*^*s*^,and *β*^*a*^ is central: asymptomatic infection is assumed to be less infectious (*β*^*a*^< *β*^*p*^), for example due to lower viral load, and *β*^*s*^ can be very low if symptomatic cases are isolated.

The basic model is described by the following equations (*i* = 1 2):

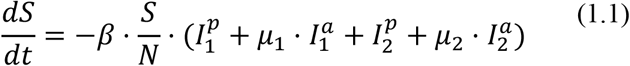

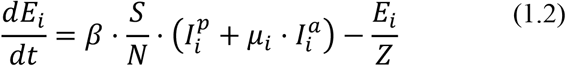

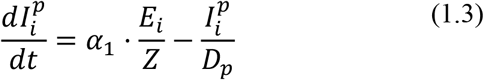

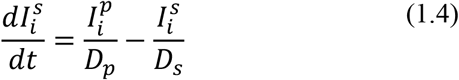

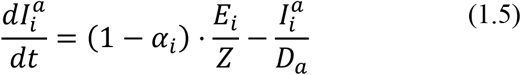

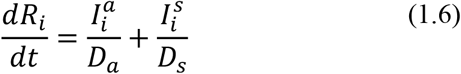

Note that 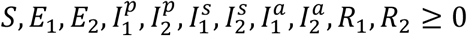 and 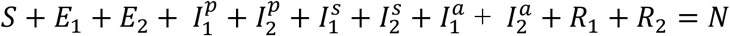, where *N* is the constant host population size.

### Basic reproduction number and stability analysis

The basic reproduction number _0_ of an epidemic can be interpreted as the expected number of secondary cases produced by a typical infected individual in a completely susceptible population^48^. It testifies for the transmissibility of the epidemic in a new host population. We define 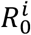, (*i* = 1,2) as the basic reproductive number for each of the viral strains in our system.

We applied the next-generation approach^49–51^ to compute the basic reproduction number. The infected compartments are 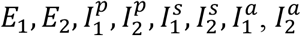. The next-generation (i.e., transition) matrix is defined as *FV*^−1^, where *F* describes the production of new infected and *V* describes transitions between infected states. The matrix has two nonzero eigenvalues, corresponding to the reproductive numbers for each strain: 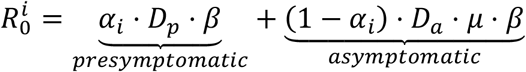 (see details in SI). Only the presymptomatic and the asymptomatic compartments contribute to *R*_0_, as individuals in the other compartments do not produce new infections. This can be interpreted as the probability that a given individual is either presymptomatic (*α*_*i*_) or asymptomatic (1 − *α*_*i*_), multiplied by the number of days from beginning of infectiousness until recovery (*D*_*p*_*D*_*a*_, for presymptomatic and asymptomatic, respectively) and the transmission rate (*β*). In the case of the test-evasive strain, the reproduction number for each strain is 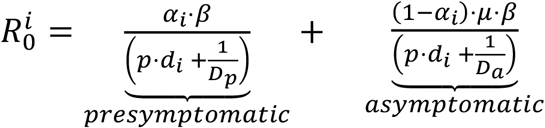 (see details in SI). The numerators are the expected numbers of secondary infections per day, and the denominators are the sums of removal rates from each infected compartment, where *p* · *d*_*i*_ is the daily detection rate.

The disease-free equilibrium (*I* ^*a*^ = *I* ^*p*^ = *I* ^*s*^ = 0) is locally unstable^52^ to the introduction of new exposed or infected individuals if 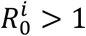 for *i* = 1 or *i* = 2. Using parameters adjusted to realistic SARS-CoV-2 values (Table 1), and specifically a transmission rate *β* ≥ 0.35, we ensure that the disease-free equilibrium in our model is locally unstable, allowing the outbreak of the epidemic for both strains.

**Table 1.** Model parameters with estimated values.

| Parameter | Description | Estimate | Source |
| --- | --- | --- | --- |
| $N$ | Total population size | 328,000,000 | US population size (2019), U.S. Census Bureau<sup>55</sup> |
| $\beta^p$ | Transmission rate in presymptomatic infected individuals | 0.35-1.2 | Li, Ruiyun, et al. 2020<sup>2</sup>. |
| $\beta^a$ | Transmission rate in asymptomatic infected individuals | $\mu \cdot \beta^p$ | Byambasuren, Oyungerel, et al. 2020<sup>33</sup>. |
| $\beta^s$ | Transmission rate in symptomatic infected individuals | 0 | Simplifying assumption that all symptomatic individuals are isolated. |
| $\alpha_1, \alpha_2$ | Fraction of presymptomatic infections | 0.6 | Oran, Daniel P., Topol, Eric J. 2021<sup>56</sup>. |
| $\mu$ | Relative infectiousness of asymptomatic infected individuals | 0.65-0.75 | Byambasuren, Oyungerel, et al. 2020<sup>33</sup>. |
| $D_p$ | Number of days in the presymptomatic phase | 3 | Casey, Miriam, et al. 2020<sup>57</sup>. |
| $D_a$ | Number of days in the asymptomatic phase | 6 | Byrne, Andrew William, et al. 2020<sup>58</sup>. |
| $D_s$ | Number of days in the symptomatic phase | 13 | Byrne, Andrew William, et al. 2020<sup>58</sup>. |
| $Z$ | Length of incubation period, or number of days in the exposed phase | 5 | McAloon, Conor, et al. 2020<sup>59</sup>. |
| $p$ | Daily tests per thousand people | 0.01-25 | Coronavirus (COVID-19) Testing - Statistics and Research - Our World in Data<sup>60</sup>. Accessed Dec. 30, 2020. |
| $d_1, d_2$ | Detectability of detectable and test-evasive strain, respectively (true positive rate) | $d_1 = 0.9$ | Arevalo-Rodriguez, Ingrid, et al. 2020<sup>41</sup>. |

### Numerical solution

To analyze the model, we use parameters estimated from COVID-19 literature (Table 1) and solve Eqs. (1.1)-(1.6) numerically using Python^53^. For the initial conditions, we assume a population of mostly susceptible individuals (*S*_0_ ∼ *N*), with a small number of exposed individuals, divided equally between the two strains (the attenuated strain can also evolve from rarity, see Fig. S1). New hosts are not introduced, so after enough time has passed, the population reaches a disease-free equilibrium (*I*^*a*^ = *I*^*p*^*= I*^*s*^ = 0). At this equilibrium, 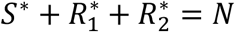, meaning all individuals are either susceptible or have recovered from the disease.

### Selection coefficient

The selection coefficient^54^ of the attenuated strain is 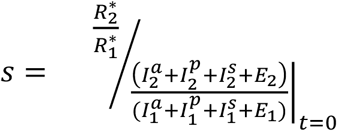, where 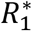 and 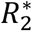 are the number of hosts at the disease-free equilibrium that have been infected by the virulent and attenuated strains, respectively, throughout the course of the epidemic. In the denominator is the ratio of initial frequencies of hosts infected by the attenuated and the virulent strain at *t* = 0. When *s* > 1, this signifies that selection favors the attenuated virus and so the frequency of the attenuated strain increases, otherwise (*s* < 1) selection favors the virulent strain. That is, the attenuated strain is expected to evolve when *s* > 1.

## Results

### Attenuation

We first consider competition between a virulent strain and an attenuated strain, where the attenuated strain has a lower fraction of symptomatic cases compared to the virulent strain. The advantage of an attenuated strain is the longer period in which individuals are infectious, as asymptomatic individuals are generally not isolated nor seek medical treatment. The disadvantage of the attenuated strain is a lower expected transmission rate (*β*), as the relative infectiousness of asymptomatic individuals is lower compared to that of presymptomatic individuals (*μ* < 1). We consider constant NPIs as a fixed reduction in the transmission rate during the entire intervention. Thus, when the impact of NPIs on transmission rates is low, the virus spreads rapidly, and the susceptible population is quickly infected by the more virulent strain. In contrast, when the impact of NPIs on transmission rates is high, the epidemic lasts longer (the curve is “flattened”), allowing the attenuated strain more time to spread.Additionally, when transmission rates are reduced, the transmission difference between the two strains is smaller. Indeed, Figure 2 shows that selection for the attenuated strain increases with the impact of NPIs and with the relative infectiousness of the attenuated strain (*μ*). We note that for a given *μ* and impact of NPI, the threshold for evolution of each strain (Fig. 2, contour line) is independent of *α*_2_, the fraction of symptomatic infected by the attenuated strain (Fig. S2).

**Figure 2.**
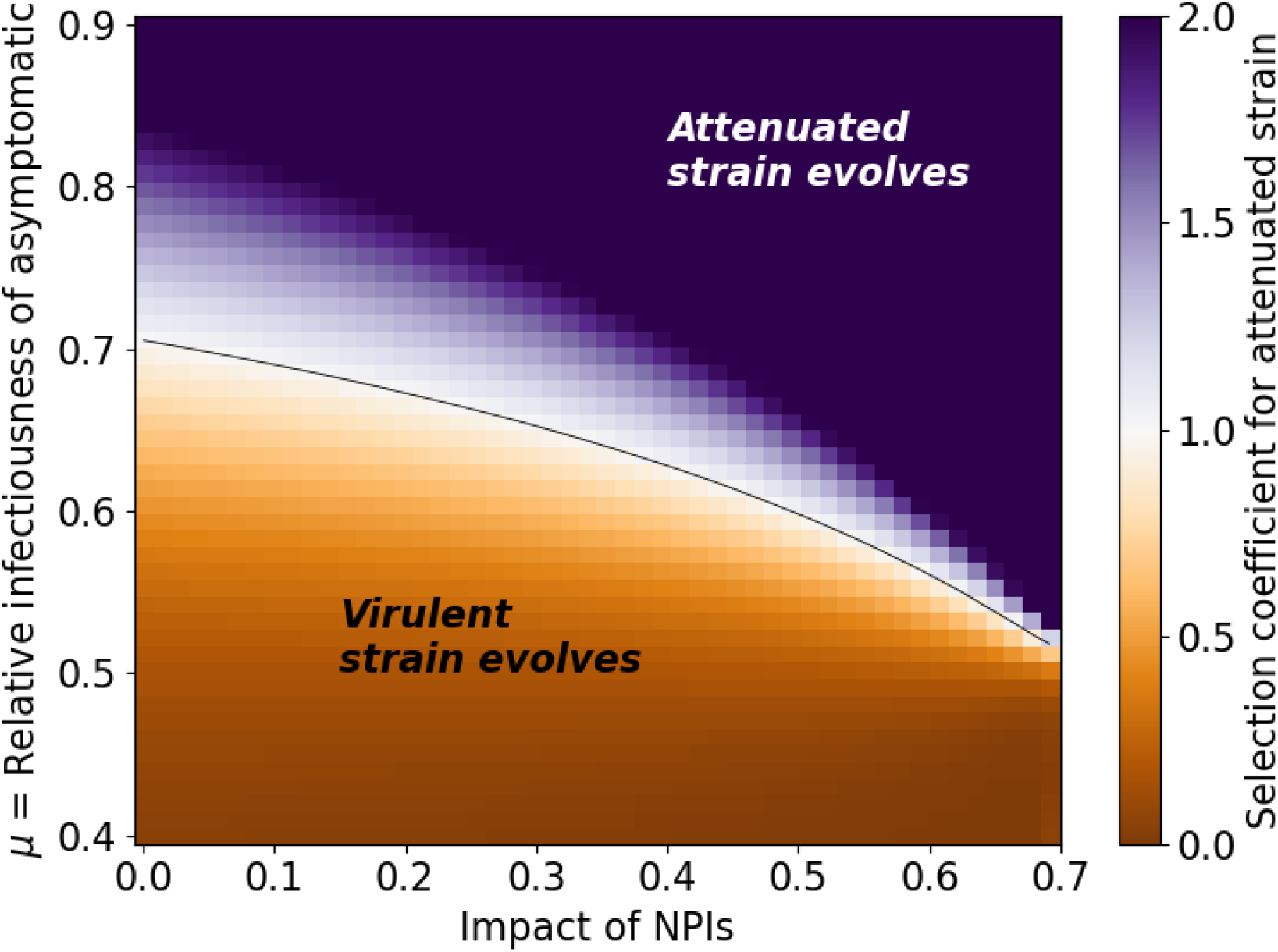
Effective NPIs facilitate the evolution of the attenuated virus. This figure presents the conditions for evolution of either the attenuated (purple) or the virulent (orange) strains under constant NPIs. High impact NPIs (right side) facilitate the evolution of the attenuated strain, while low impact NPIs (left side) facilitate the evolution of the virulent strain. The attenuated strain can also evolve if asymptomatic individuals are infectious enough relative to presymptomatic individuals (*μ* is high, top side). Here, *β*_max_= 1.2,*α*_1_ = 0.6, *α*_2_ = 0.05.

NPIs have been implemented with various schedules, determined by epidemiological metrics^61^, economic pressures^61^, public opinion^62^, and in some cases have probably started later than planned^63^. Next, we explore the effects of temporal application of NPIs^7,64^ on the competition between attenuated and virulent strains. Figure 3 shows the effect of the number of days between the outbreak and start of NPIs on the competition between the two viral strains. Overall, if NPIs are implemented earlier, then the attenuated strain is more likely to evolve. As above (Figure 2), lower impact NPIs favor the virulent strain, while higher impact NPIs favor the attenuated strain.

**Figure 3.**
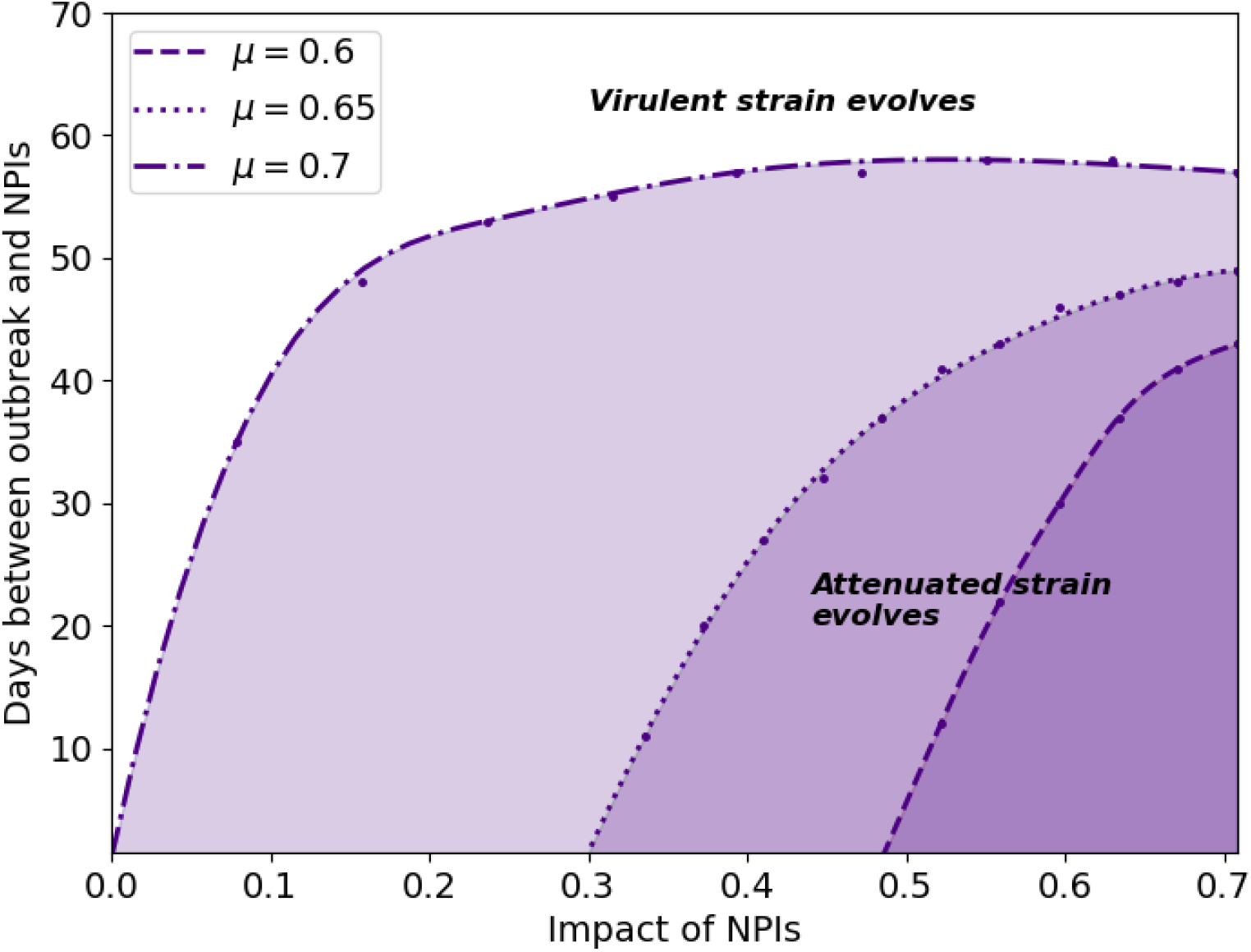
Earlier start of NPIs favors the evolution of the attenuated strain. Each curve corresponds to a different relative transmissibility of infection by asymptomatic individuals, *μ*. The colored areas below each curve show the regions in which the attenuated strain evolves. The areas above each of the curves show the regions in which the virulent strain evolves. Here, *β*_min_= 0.35, *β*_max_= 1.2,*α*_1_ = 0.6, *α*_2_ = 0.05.

Figure 4 compares the dynamics without NPIs (left) and with NPIs that begin a certain number of days after the outbreak and are lifted after a limited time (right). We find that NPIs significantly affect the competition between the two strains, changing the direction of selection on the virus and leading to the evolution of the attenuated strain (compare panels *a* and *b*). While NPIs reduce the peak number of symptomatic cases (compare panels *c* and *d*), a “second wave” of infections may occur when NPIs are over (panels *d* and *f*). Here, this “second wave” is dominated by the attenuated strain (panel *f*) that produces more asymptomatic infections compared to the virulent strain (compare purple dashed line and orange solid line in panel *d*).

**Figure 4.**
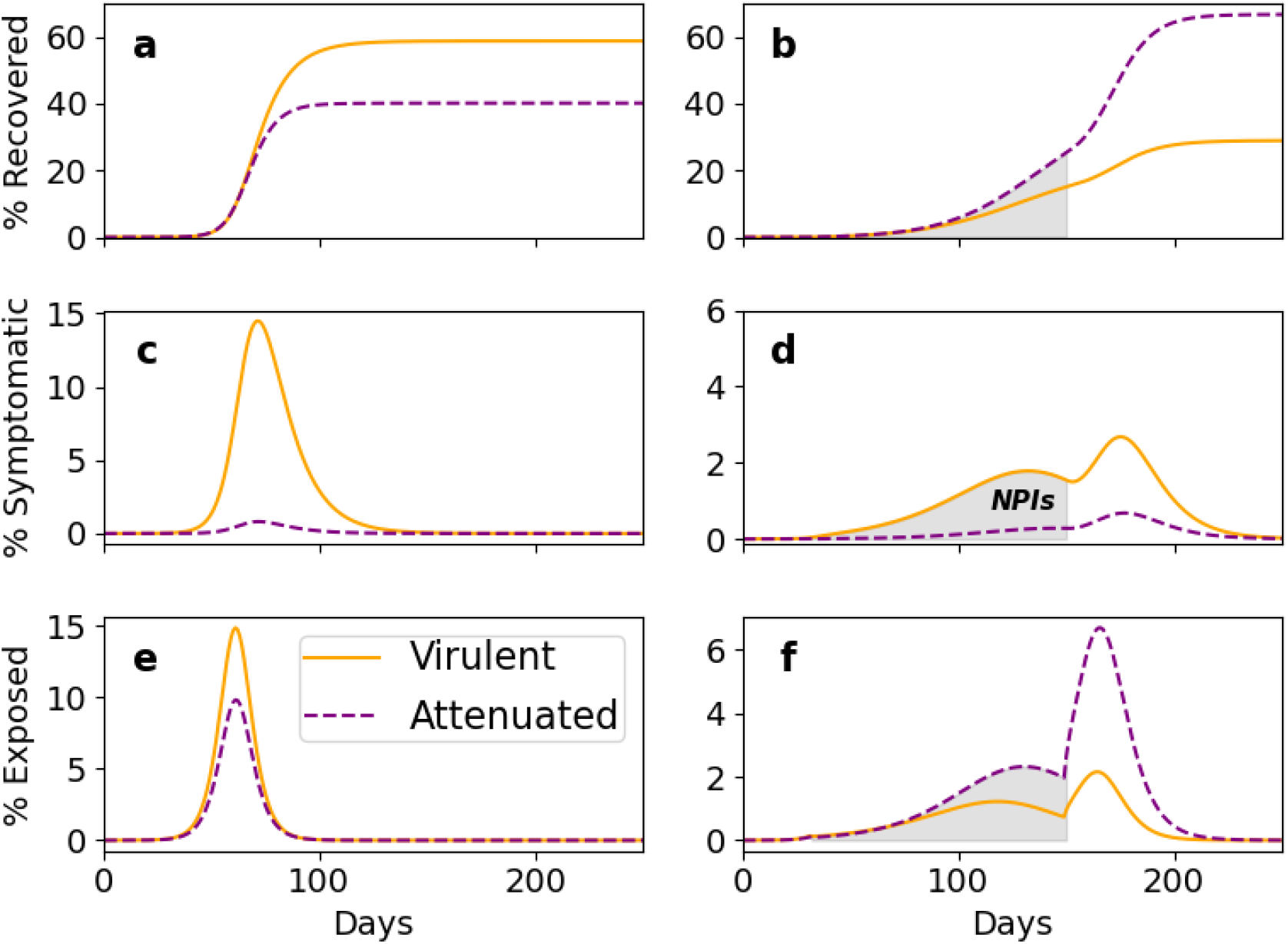
Temporal application of NPIs favors the evolution of the attenuated strain. Without NPIs (left), the virulent strain takes over the viral population. When NPIs are applied (right, shaded area), the virulent strain is more frequent among symptomatic (panel *d*), but the attenuated strain becomes more frequent in exposed individuals during the NPIs (panel *f*), and after the NPIs end it is the dominant strain (panel *b*). Parameters: NPIs start on day 31 and end on day 150. *μ* = 0.6,*β*_min_= 0.42,*β*_max_= 1.2.

### Test evasion

We assume that a proportion *p* of the population is tested for SARS-CoV-2 infection every day and consider the evolution of a test-evasive strain. *Detectability* is defined here as the test sensitivity, or the true positive rate: the probability that an infected individual will be correctly detected by a single test. The detectability of the detectable and test-evasive strains is *d*_1_,*d*_2_ respectively, where *d*_2_ < *d*_1_. All else being equal, the test-evasive strain will evolve due to its lower detectability (Fig. S3a). However, lower detectability likely incurs a cost for the virus, as the target sequences for SARS-CoV-2 tests are in regions essential for replication and other critical aspects of the viral life cycle^39^. We assume this cost, *c*,is expressed in decreased infectiousness, such that *β*_2_ = (1 −*c*) *β*_1_ (see Eq. S2.1 in supplementary). Thus, we examine a competition between a detectable and a test-evasive strain that is less infectious compared to the virulent strain, *c* > 0.

Figure 5 shows that a higher testing rate (*p*) may select for a test-evasive strain, even when reduced detectability incurs decreased infectiousness. When the impact of NPIs is higher (right), the test-evasive strain evolves even when the testing rate is low (solid line).

**Figure 5.**
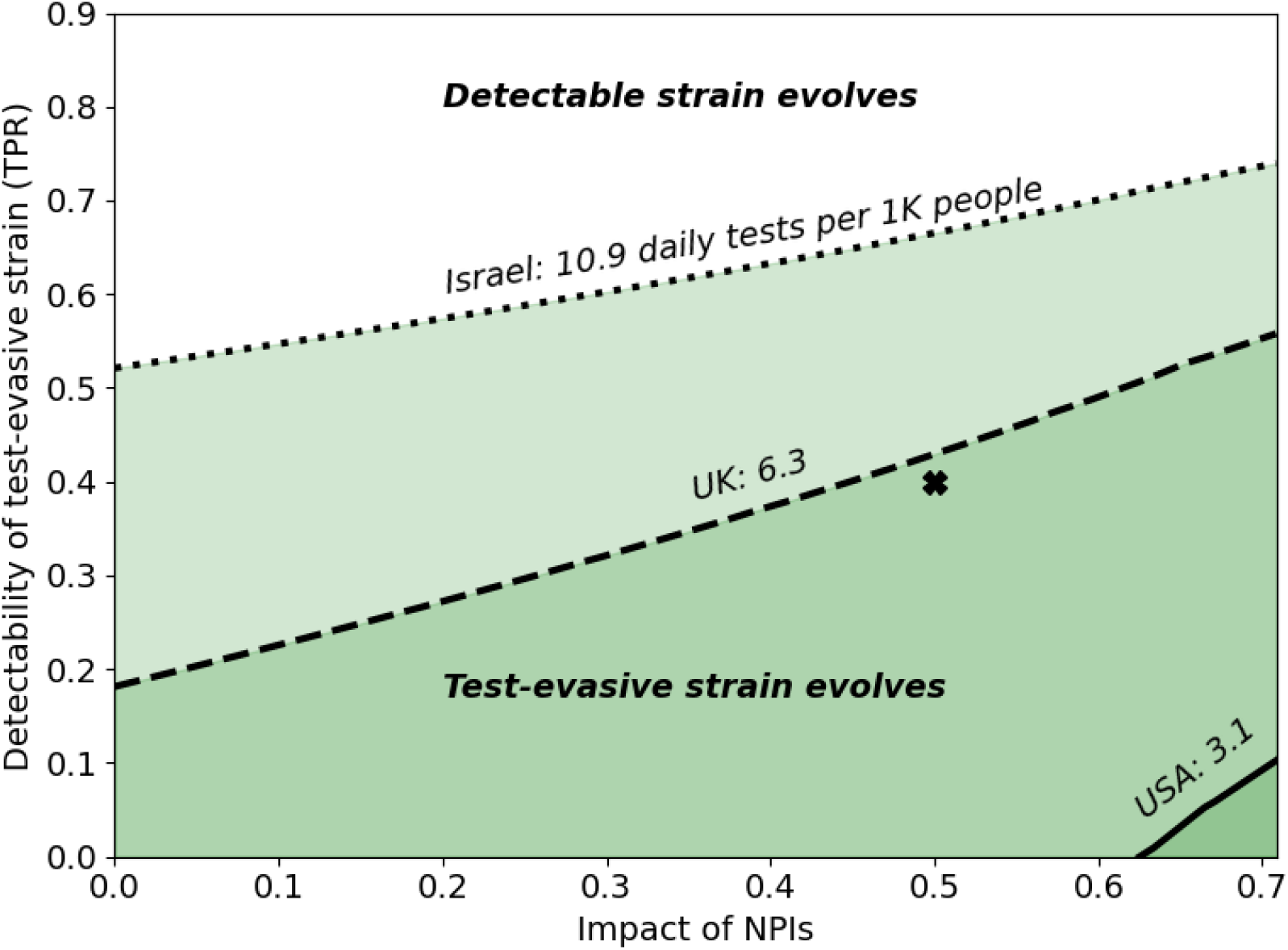
High testing rate and effective NPIs favor the evolution of test-evasive strains. This figure presents the conditions for the evolution of test-evasive strains despite a cost of infectiousness (see Fig, S3b for further decreased infectiousness). Each line corresponds to a different testing rate, *p*. The colored areas below each line show the regions in which the test-evasive strain evolves. The areas above each of the lines show the regions in which the detectable strain evolves. Test-evasive strains can also evolve if the detectability of the test-evasive strain is low enough. For example, given that the impact of NPIs is 0.5, a test-evasive strain with detectability *d*_2_ = 0.4(marked here with ‘X’) would evolve in Israel and the UK, but in the USA the more detectable strain would evolve. Here, *α*_1_ = 0.6,*α*_2_ = 0.6,*μ* = 0.6,*c* = 0.01,*d*_1_ = 0.9. Estimated values for testing rates in different countries were accessed on December 30, 2020 (Table 1).

In Figure 6, we examine the effects of decreased detectability on epidemiological metrics. We explore two scenarios of epidemic outbreaks: exclusively by a detectable strain (black lines), and exclusively by a test-evasive strain (green lines). The number of confirmed daily cases increases with the testing rate (Fig. 6c, compare solid, dashed, and dotted lines), while the number of actual daily cases decreases with it (Fig. 6a, compare solid, dashed and dotted lines). When the epidemic is driven by a test-evasive strain (green lines), the daily number of confirmed cases and daily percent of positive tests (Fig. S5) increase with the detectability of the test-evasive strain (Fig. 6c), while the daily number of actual cases decreases (Fig. 6a). Overall, when testing rate is high and the epidemic is driven by a test-evasive strain with low detectability (Figs. 6b and 6d), the number of confirmed cases can be misleading: it is significantly lower for the test-evasive strain, despite the number of actual cases being higher.

**Figure 6.**
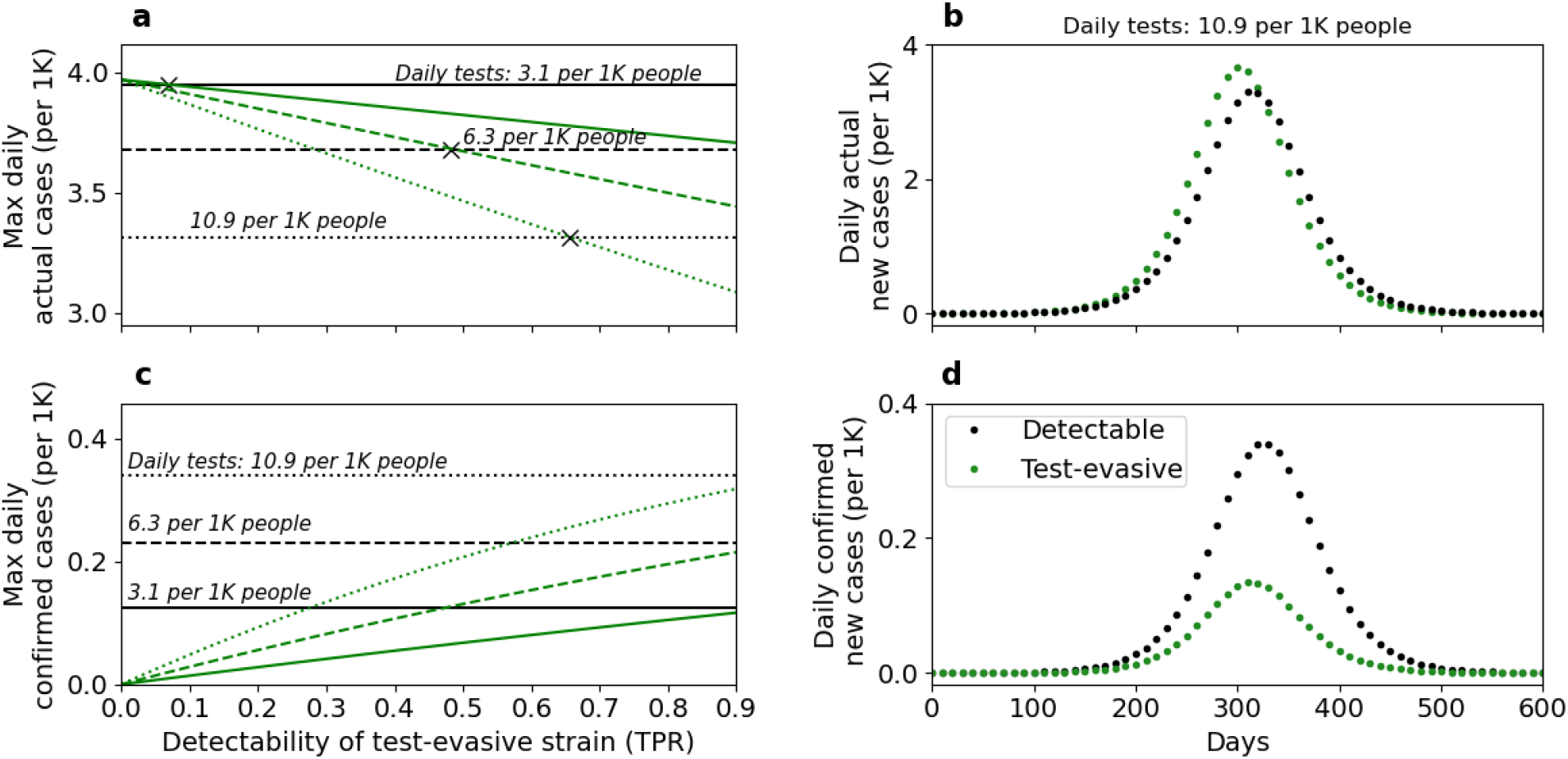
Effects of decreased detectability on epidemiological metrics. These results demonstrate the outcomes of two separate outbreaks: (i) exclusively by a detectable strain (black lines) and (ii) exclusively by a test-evasive strain (green lines), under three testing regimes (solid, dashed, and dotted lines). Panels ***a*** and ***c*** show the maximum number of actual and test-confirmed daily cases, respectively. Panels ***b*** and ***d*** show a timeline of these epidemiological metrics for a relatively high testing rate and low detectability. Higher detectability of the test-evasive strain increases the daily number of confirmed cases (panel ***c***, green lines) and decreases the daily number of actual cases (panel ***a***, green lines). The effect of detectability on the number of daily actual cases is associated with the cost of infectiousness, *c*, producing a detectability threshold (panel ***a***, marked with ‘X’), above which the number of actual daily cases for the test-evasive strain is lower than for the detectable strain (see Fig. S4 for the effect of decreased detectability without cost of infectiousness). A higher testing rate increases the number of confirmed cases (panel **c**, compare solid, dashed and dotted lines) and decreases the number of actual cases (panel ***a***, compare solid, dashed and dotted lines). Overall, when testing rate is high and the epidemic is driven by a test-evasive strain with low detectability (panels ***b*** and ***d***), the number of confirmed cases is lower compared to an epidemic driven by a detectable strain, while the number of actual cases is higher. Here, *β* = 0.42 (Impact of NPI = 0.65), *μ* = 0.6,*α*_1_ = 0.6,*α*_2_ = 0.6,*d*_1_ = 0.9,*c* = 0.01. For panels ***b*** and ***d***, *d*_2_ = 0.3,*p* = 10.9 per 1K people.

## Discussion

We examined the expected selection pressures exerted by non-pharmaceutical interventions and testing on virulence and detectability of SARS-CoV-2. We found that when stronger NPIs are applied, selection may act to reduce virulence. Additionally, the timely application of NPIs could significantly affect the competition between viral strains, favoring reduced virulence. Furthermore, a higher testing rate can select for a test-evasive viral strain, even if that strain is less infectious than the competing detectable strain. Our results also show that if a test-evasive strain evolves, reductions in epidemiological metrics such as confirmed daily cases may be due to reductions in test sensitivity rather than reductions in the actual number of cases.

Our model makes several simplifying assumptions. We assume that individuals exhibiting clinical symptoms are isolated and therefore do not transmit the virus (*β*_*c*_ = 0). However, such individuals may still be able to infect others, whether it is in a medical facility, within the household, or due to non-compliance with isolation guidelines. Nevertheless, practical strategies have been put in place to reduce nosocomial transmission^65^, and evidence suggests that the overall risk of hospital-acquired COVID-19 is low^65^. In our model, when the impact of NPIs is low, the susceptible population is infected rapidly by the more virulent strain. Our model could be extended by allowing individuals that recovered from one strain to become infected with another strain, and in that case the attenuated strain may evolve even when the impact of NPIs is low. We assume that the entire population is tested daily with a testing rate estimated by realistic parameters (Figs. 5 and 6). However, the average testing rate likely underestimates the rate for infected cohorts, as individuals who experience symptoms or have been exposed to a confirmed case are more inclined to be tested. Applying a higher testing rate would make the evolution of the test-evasive strain even more likely (Fig. 5).

The recent emergence of novel SARS-CoV-2 variants has raised widespread concern^26,27^. SARS-CoV-2 is likely a pathogen of recent zoonotic origin, and can therefore further adapt to its new human host^66^. Some existing mutations have been said to increase infectiousness^67^ and be associated with a younger patient age^68^. Some variants are suspected of ‘immune escape’, eluding the human immune response^69,70^, such that more recovered individuals remain susceptible to reinfection and possibly causing a reduction in the effectiveness of vaccines^69,70^. The future evolutionary and epidemiological trajectories of the virus are difficult to predict^71,72^, and it may evolve into different variants differing in their level of virulence and transmissibility^66^. Our results show that non-pharmaceutical interventions and testing policies, primarily designed and applied to control the spread of the pandemic, may also steer the evolution of the virus towards attenuation and test-evasion.

## Data Availability

The code used to produce the results is available upon request.

## Acknowledgements and funding

This project was supported by Israel Science Foundation grant 3811/19 (L.H. and Y.R.) and Minerva Stiftung Center for Lab Evolution (L.H. and Y.R.). We thank Uri Obolski for discussions.

## Author Contributions

Y.G., Y.R., and L.H. designed the study and formulated the model. Y.G. derived the equations and implemented the code. Y.G., Y.R., and L.H. analysed the results and wrote the manuscript.

