## Supplementary Information for "Modeling the evolution of SARS-CoV-2 under non-pharmaceutical interventions"

#### Attenuation

The basic model is described by

$$\frac{dS}{dt} = -\beta \cdot \frac{S}{N} \cdot (I_1^p + \mu_1 \cdot I_1^a + I_2^p + \mu_2 \cdot I_2^a) \tag{S1.1}$$

$$\frac{dE_i}{dt} = \beta \cdot \frac{S}{N} \cdot \left(I_i^p + \mu_i \cdot I_i^a\right) - \frac{E_i}{Z} \tag{S1.2}$$

$$\frac{dI_i^p}{dt} = \alpha_i \cdot \frac{E_i}{Z} - \frac{I_i^p}{D_p} \tag{S1.3}$$

$$\frac{dI_i^s}{dt} = \frac{I_i^p}{D_p} - \frac{I_i^s}{D_s} \tag{S1.4}$$

$$\frac{dI_i^a}{dt} = (1 - \alpha_i) \cdot \frac{E_i}{Z} - \frac{I_i^a}{D_a} \tag{S1.5}$$

$$\frac{dR_i}{dt} = \frac{I_i^a}{D_a} + \frac{I_i^s}{D_s} \tag{S1.6}$$

where subscript $i$ stands for the strains $i = 1{,}2$ and superscript *p*, *s*, and *a* stand for presymptomatic, symptomatic, and asymptomatic infections. We assume that individuals exhibiting clinical symptoms ($I^s$) are isolated and therefore do not transmit the virus.

**Basic reproduction number.** We applied the next-generation approach[1,2] to compute the basic reproduction number of the epidemic. The infected compartments are $E_1, I_1^p, I_1^s, I_1^a, E_2, I_2^p, I_2^s, I_2^a$. The next-generation (i.e., transition) matrix is defined as $FV^{-1}$, where $F$ describes the production of new infected and $V$ describes transitions between infected states.

$F\ =$

| | $\boldsymbol{E_1}$ | $\boldsymbol{I_1^p}$ | $\boldsymbol{I_1^s}$ | $\boldsymbol{I_1^a}$ | $\boldsymbol{E_2}$ | $\boldsymbol{I_2^p}$ | $I_2^S$ | $\boldsymbol{I_2^a}$ |
|---|---|---|---|---|---|---|---|---|
| $\boldsymbol{E_1}$ | 0 | $\beta \cdot \frac{S}{N}$ | 0 | $\beta \cdot \mu \cdot \frac{S}{N}$ | 0 | 0 | 0 | 0 |
| $\boldsymbol{I_1^p}$ | 0 | 0 | 0 | 0 | 0 | 0 | 0 | 0 |
| $\boldsymbol{I_1^s}$ | 0 | 0 | 0 | 0 | 0 | 0 | 0 | 0 |
| $\boldsymbol{I_1^a}$ | 0 | 0 | 0 | 0 | 0 | 0 | 0 | 0 |
| $\boldsymbol{E_2}$ | 0 | 0 | 0 | 0 | 0 | $\beta \cdot \frac{S}{N}$ | 0 | $\beta \cdot \mu \cdot \frac{S}{N}$ |
| $\boldsymbol{I_2^p}$ | 0 | 0 | 0 | 0 | 0 | 0 | 0 | 0 |
| $\boldsymbol{I_2^s}$ | 0 | 0 | 0 | 0 | 0 | 0 | 0 | 0 |
| $\boldsymbol{I_2^a}$ | 0 | 0 | 0 | 0 | 0 | 0 | 0 | 0 |

$V\ =$

| | $\boldsymbol{E_1}$ | $\boldsymbol{I_1^p}$ | $\boldsymbol{I_1^s}$ | $\boldsymbol{I_1^a}$ | $\boldsymbol{E_2}$ | $\boldsymbol{I_2^p}$ | $I_2^S$ | $\boldsymbol{I_2^a}$ |
|---|---|---|---|---|---|---|---|---|
| $\boldsymbol{E_1}$ | $\frac{1}{Z}$ | 0 | 0 | 0 | 0 | 0 | 0 | 0 |
| $\boldsymbol{I_1^p}$ | $-\frac{\alpha_1}{Z}$ | $\frac{1}{D_p}$ | 0 | 0 | 0 | 0 | 0 | 0 |
| $\boldsymbol{I_1^s}$ | 0 | $-\frac{1}{D_p}$ | $\frac{1}{D_s}$ | 0 | 0 | 0 | 0 | 0 |
| $\boldsymbol{I_1^a}$ | $-\frac{(1-\alpha_{1)}}{Z}$ | 0 | 0 | $\frac{1}{D_a}$ | 0 | 0 | 0 | 0 |
| $\boldsymbol{E_2}$ | 0 | 0 | 0 | 0 | $\frac{1}{Z}$ | 0 | 0 | 0 |
| $\boldsymbol{I_2^p}$ | 0 | 0 | 0 | 0 | $-\frac{\alpha_2}{Z}$ | $\frac{1}{D_p}$ | 0 | 0 |
| $I_2^S$ | 0 | 0 | 0 | 0 | 0 | $-\frac{1}{D_p}$ | $\frac{1}{D_s}$ | 0 |
| $\boldsymbol{I_2^a}$ | 0 | 0 | 0 | 0 | $-\frac{(1-\alpha_{2)}}{Z}$ | 0 | 0 | $\frac{1}{D_a}$ |

We used Matlab R2020b (*eig* function) to find the eigenvalues of the matrix $FV^{-1}$. The matrix has two nonzero eigenvalues, corresponding to the reproductive numbers for each strain: $R_0^i = \underbrace{\alpha_i \cdot D_p \cdot \beta}_{presymptomatic} + \underbrace{(1-\alpha_i) \cdot D_a \cdot \mu \cdot \beta}_{asymptomatic}$.

**Initial conditions.** We initialize the epidemic timeline at $t = 0$ with $I_1^a = I_2^a = 750$, $E_1 = E_2 = 250$. We examined the effect of initial conditions (Fig. S1) to show that the initial conditions do not have a significant effect on the evolution of the attenuated strain.

**Impact of NPIs.** Throughout the main text, we use the term "Impact of NPIs" (x-axis in Fig. 2, Fig. 3, Fig. 5). It is defined as $1 - \frac{\beta_{min}}{\beta_{max}}$, where $\beta_{min}$ and $\beta_{max}$ are given in the figure captions.

**Equilibria.** Our system cannot reach an endemic equilibrium (steady positive $E$ and/or $I$ compartments) because we neglect host recruitment ($\frac{dS}{dt} \leq 0$ always). To compute the disease-free equilibrium ($E = I = 0$), we set the rate of change of all state variables to zero. Solving the system of algebraic equations, we find a disease-free equilibrium,

$$\left(S^*, E_1^*, E_2^*, I_1^{p*}, I_2^{p*}, I_1^{s*}, I_2^{s*}, I_1^{a*}, I_2^{a*}, R_1^*, R_2^*\right) = (S^*, 0,0,0,0,0,0,0,0,0, N - S^*),$$

where $S^*$, the number of susceptible individuals at equilibrium, can take values within the range $0 \leq S^* \leq N$ depending on the initial conditions and disease dynamics.

#### Test-evasion

We add an additional compartment, $Q$ for quarantined, for infected individuals that are isolated after receiving a positive test result. We note that symptomatic cases ($I^s$) are isolated due to clinical symptoms, regardless of testing. Quarantined individuals ($Q$) are treated as if they are symptomatic, hence leave quarantine after $D_s$ days. We assume the test-evasive strain incurs a cost of infectiousness, $c > 0$, such that $\beta_2 = (1 - c) \cdot \beta_1$. We assume a proportion $p$ of the population (except for the $Q$ compartment) is tested daily and that $d_1, d_2$ are the sensitivities (true positive rates) of the test for the detectable and the test-evasive strain, respectively. We neglect false-positive results, assuming 100% test specificity (FPR=0). Exposed ($E$) and infected ($I^s, I^a, I^p$) compartments are likely tested after exposure to an infected or due to symptoms (mild for $I^p$, more severe for $I^s$). However, we neglect early detection and assume only infected may test positive, as most of those exposed individuals test negative[3]. Susceptible individuals ($S$) may also be tested due to exposure to infecteds, or due to symptoms similar to but independent of COVID-19. Susceptible individuals may also be tested to obtain a clean bill of health (e.g., for travel purposes) or as part of a testing effort for individuals not exhibiting symptoms. Individuals in the removed compartment ($R$) are also tested, because they may not be aware they were previously infected.

The model with testing is described by (changed from Eq. S1 are in bold)

$$\frac{dS}{dt} = -\beta \cdot \frac{S}{N} \cdot \left(I_1^p + \mu_1 \cdot I_1^a\right) - \beta \cdot \frac{S}{N} \cdot (\mathbf{1} - \boldsymbol{c}) \cdot (I_2^p + \mu_2 \cdot I_2^a) \quad \text{(S2.1)}$$

$$\frac{dE_i}{dt} = \beta \cdot \frac{S}{N} \cdot \left(I_i^p + \mu_i \cdot I_i^a\right) - \frac{E_i}{Z} \quad \text{(S2.2)}$$

$$\frac{dI_i^p}{dt} = \alpha_1 \cdot \frac{E_i}{Z} - \frac{I_i^p}{D_p} - \boldsymbol{p} \cdot \boldsymbol{d_i} \cdot \boldsymbol{I_i^p} \quad \text{(S2.3)}$$

$$\frac{dI_i^s}{dt} = \frac{I_i^p}{D_p} - \frac{I_i^s}{D_s} - \boldsymbol{p} \cdot \boldsymbol{d_i} \cdot \boldsymbol{I_i^p} \quad \text{(S2.4)}$$

$$\frac{dI_i^a}{dt} = (1 - \alpha_i) \cdot \frac{E_i}{Z} - \frac{I_i^a}{D_a} - \boldsymbol{p} \cdot \boldsymbol{d_i} \cdot \boldsymbol{I_i^a} \quad \text{(S2.5)}$$

$$\boldsymbol{\frac{dQ_i}{dt}} = \boldsymbol{p} \cdot \boldsymbol{d_i} \cdot (\boldsymbol{I_i^a} + \boldsymbol{I_i^p} + \boldsymbol{I_i^s}) - \boldsymbol{\frac{Q_i}{D_s}} \quad \text{(S2.6)}$$

$$\frac{dR_i}{dt} = \frac{I_i^a}{D_a} + \frac{I_i^s}{D_s} + \boldsymbol{\frac{Q_i}{D_s}} \quad \text{(S2.7)}$$

**Epidemiological metrics** used in Fig. 6 in the main text, and Fig. S4 and Fig. S5 in the SI are

| | |
|---|---|
| **Daily actual cases** | $\frac{E_1}{Z}+\frac{E_2}{Z}$ |
| **Daily confirmed cases** | $p \cdot d_1 \cdot (I_1^a + I_1^p + I_1^s) + p \cdot d_2 \cdot (I_2^a + I_2^p + I_2^s)$ |
| **Daily % positive tests** | $\frac{p \cdot d_1 \cdot (I_1^a + I_1^p + I_1^s) + p \cdot d_2 \cdot (I_2^a + I_2^p + I_2^s)}{p \cdot (N - Q_1 - Q_2)}$ |

**Basic reproduction number with testing.** We computed the basic reproduction number of the epidemic using the next-generation approach similarly to the attenuation model. The infected compartments are $E_1, I_1^p, I_1^s, I_1^a, Q_1, E_2, I_2^p, I_2^s, I_2^a, Q_2$. The next-generation (i.e., transition) matrix is defined as $FV^{-1}$, where $F$ describes the production of new infected and $V$ describes transitions between infected states.

$F =$

| | $\boldsymbol{E_1}$ | $\boldsymbol{I_1^p}$ | $\boldsymbol{I_1^s}$ | $\boldsymbol{I_1^a}$ | $\boldsymbol{Q_1}$ | $\boldsymbol{E_2}$ | $\boldsymbol{I_2^p}$ | $\boldsymbol{I_2^s}$ | $\boldsymbol{I_2^a}$ | $\boldsymbol{Q_2}$ |
|---|---|---|---|---|---|---|---|---|---|---|
| $\boldsymbol{E_1}$ | 0 | $\beta \cdot \frac{S}{N}$ | 0 | $\beta \cdot \mu \cdot \frac{S}{N}$ | 0 | 0 | 0 | 0 | 0 | 0 |
| $\boldsymbol{I_1^p}$ | 0 | 0 | 0 | 0 | 0 | 0 | 0 | 0 | 0 | 0 |
| $\boldsymbol{I_1^s}$ | 0 | 0 | 0 | 0 | 0 | 0 | 0 | 0 | 0 | 0 |
| $\boldsymbol{I_1^a}$ | 0 | 0 | 0 | 0 | 0 | 0 | 0 | 0 | 0 | 0 |
| $\boldsymbol{Q_1}$ | 0 | 0 | 0 | 0 | 0 | 0 | 0 | 0 | 0 | 0 |
| $\boldsymbol{E_2}$ | 0 | 0 | 0 | 0 | 0 | 0 | $\beta \cdot \frac{S}{N}$ | 0 | $\beta \cdot \mu \cdot \frac{S}{N}$ | 0 |
| $\boldsymbol{I_2^p}$ | 0 | 0 | 0 | 0 | 0 | 0 | 0 | 0 | 0 | 0 |
| $\boldsymbol{I_2^s}$ | 0 | 0 | 0 | 0 | 0 | 0 | 0 | 0 | 0 | 0 |
| $\boldsymbol{I_2^a}$ | 0 | 0 | 0 | 0 | 0 | 0 | 0 | 0 | 0 | 0 |
| $\boldsymbol{Q_2}$ | 0 | 0 | 0 | 0 | 0 | 0 | 0 | 0 | 0 | 0 |

$V\ =$

| | $\boldsymbol{E_1}$ | $\boldsymbol{I_1^p}$ | $\boldsymbol{I_1^s}$ | $\boldsymbol{I_1^a}$ | $\boldsymbol{Q_1}$ | $\boldsymbol{E_2}$ | $\boldsymbol{I_2^p}$ | $\boldsymbol{I_2^s}$ | $\boldsymbol{I_2^a}$ | $\boldsymbol{Q_2}$ |
|---|---|---|---|---|---|---|---|---|---|---|
| $\boldsymbol{E_1}$ | $\frac{1}{Z}$ | 0 | 0 | 0 | 0 | 0 | 0 | 0 | 0 | 0 |
| $\boldsymbol{I_1^p}$ | $-\frac{\alpha_1}{Z}$ | $\frac{1}{D_p}+p\cdot d_1$ | 0 | 0 | 0 | 0 | 0 | 0 | 0 | 0 |
| $\boldsymbol{I_1^s}$ | 0 | $-\frac{1}{D_p}$ | $\frac{1}{D_s}+p\cdot d_1$ | 0 | 0 | 0 | 0 | 0 | 0 | 0 |
| $\boldsymbol{I_1^a}$ | $-\frac{(1-\alpha_1)}{Z}$ | 0 | 0 | $\frac{1}{D_a}+p\cdot d_1$ | 0 | 0 | 0 | 0 | 0 | 0 |
| $\boldsymbol{Q_1}$ | 0 | $-p\cdot d_1$ | $-p\cdot d_1$ | $-p\cdot d_1$ | $\frac{1}{D_s}$ | 0 | 0 | 0 | 0 | 0 |
| $\boldsymbol{E_2}$ | 0 | 0 | 0 | 0 | 0 | $\frac{1}{Z}$ | 0 | 0 | 0 | 0 |
| $\boldsymbol{I_2^p}$ | 0 | 0 | 0 | 0 | 0 | $-\frac{\alpha_2}{Z}$ | $\frac{1}{D_p}+p\cdot d_2$ | 0 | 0 | 0 |
| $\boldsymbol{I_2^s}$ | 0 | 0 | 0 | 0 | 0 | 0 | $-\frac{1}{D_p}$ | $\frac{1}{D_s}+p\cdot d_2$ | 0 | 0 |
| $\boldsymbol{I_2^a}$ | 0 | 0 | 0 | 0 | 0 | $-\frac{(1-\alpha_2)}{Z}$ | 0 | 0 | $\frac{1}{D_a}+p\cdot d_2$ | 0 |
| $\boldsymbol{Q_2}$ | 0 | 0 | 0 | 0 | 0 | 0 | $-p\cdot d_2$ | $-p\cdot d_2$ | $-p\cdot d_2$ | $\frac{1}{D_s}$ |

We used Matlab R2020b (*eig* function) to find the eigenvalues of the matrix $FV^{-1}$. The matrix has two nonzero eigenvalues, corresponding to the reproductive numbers for each strain: $R_0^i =$

$$\underbrace{\frac{\alpha_i \cdot \beta}{\left(p \cdot d_i + \frac{1}{D_p}\right)}}_{presymptomatic} + \underbrace{\frac{(1-\alpha_i) \cdot \mu \cdot \beta}{\left(p \cdot d_i + \frac{1}{D_a}\right)}}_{asymptomatic} .$$

See parameter Table 1 in the main text. The numerators are the expected number of secondary infections per day. The denominators are the sums of removal rates from each compartment, where $p \cdot d_i$ is the daily detection rate.

### Supplementary Figures

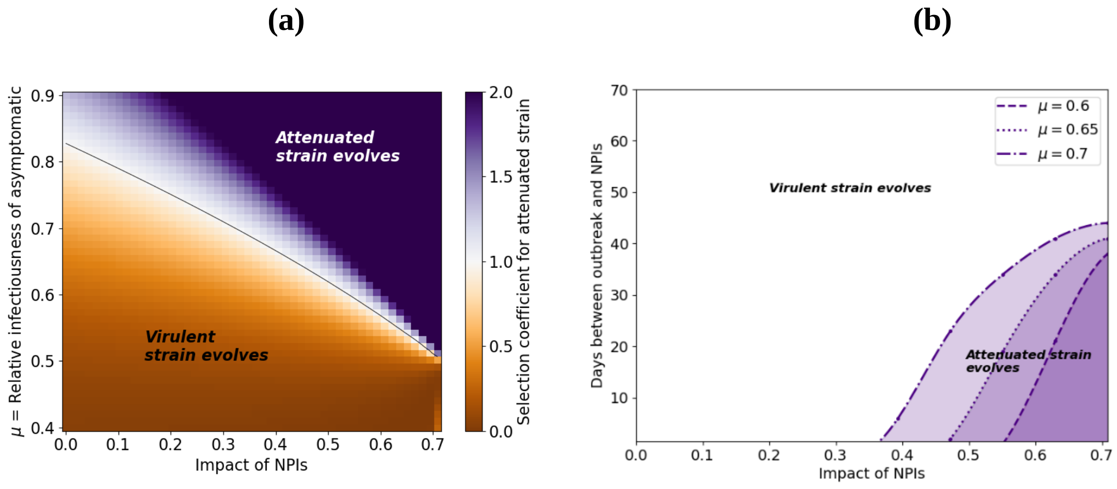


**Figure S1: Effective NPIs allow for the evolution of the attenuated virus from rarity.** The evolution of the attenuated strain is defined by its frequency increasing from the initial frequency. In the main text, we set the number of individuals initially infected with each of the strains to be equal (infected with attenuated strain are 50% of total infected). Here we examine the evolution of the attenuated strain when the initial number of individuals infected with the attenuated strain is 0.05% of the total infected. **(a)** These results demonstrate that the attenuated strain can evolve from rarity under a wide range of conditions. **(b)** To allow the evolution of the attenuated strain from rarity, stronger and earlier NPIs should be applied (compare to Fig. 3 in the main text). Here, $\beta_{\max} = 1.2,\ \alpha_1 = 0.6, \alpha_2 = 0.05$. Initial conditions: $I_1^a = 1500,\ I_2^a = 0$ , $E_1 = 499, E_2 = 1$.

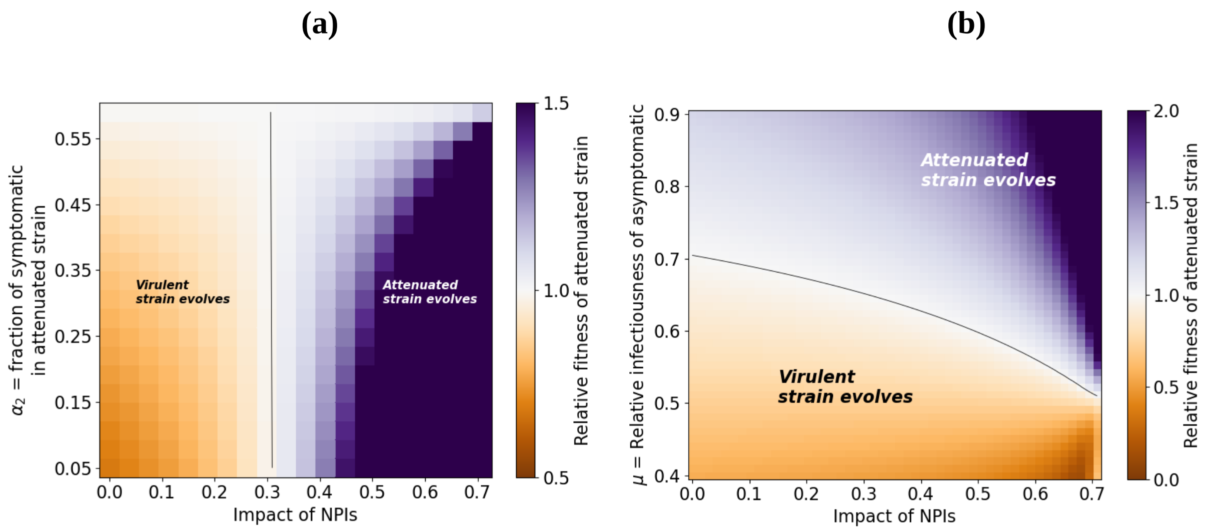


**Figure S2: The effect of symptomatic rate ($\alpha_2$) on the evolution of the attenuated strain. (a)** For a given $\mu$ (relative infectiousness of the attenuated strain), if the impact of NPIs is such that the virulent strain would evolve (Fig. S2a, orange), the relative fitness of the attenuated strain increases with $\alpha_2$ (larger fraction of symptomatic). However, if the impact of NPIs is such that attenuated strain would evolve (Fig. S2a, purple), the relative fitness of the attenuated strain decreases with $\alpha_2$. Nevertheless, these results show that for a given $\mu$, the threshold for the evolution of the attenuated strain (contour line) is independent of $\alpha_2$ and depends only on the impact of NPIs. **(b)** Evolution of attenuated strain with $\alpha_2 = 0.5$. Comparing to Fig. 2 in the main text ($\alpha_2 = 0.05$), the relative fitness of the attenuated strain decreases while the relative fitness of the virulent strain increases throughout the range of parameters. However, the threshold of evolution for the attenuated strain does not change (contour line). Here, (a) $\mu = 0.6$. (b) $\alpha_2 = 0.5$.

**(a)** **(b)**

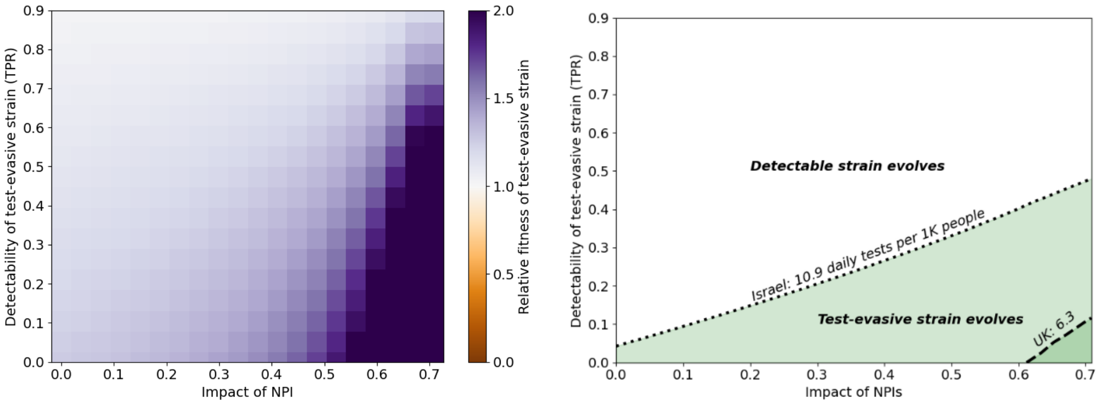


**Figure S3: The effect of the cost of infectiousness on the evolution of the test-evasive strain.** **(a)** Without a cost of infectiousness, and with all else being equal, the test-evasive strain will always evolve. **(b)** Increased testing rate selects for test-evasive strains despite an increased cost of infectiousness. Effective NPIs and low detectability select for test-evasive strains. However, here we further increase the cost of infectiousness (compare to Fig. 5 in main text) and show that a higher cost of infectiousness reduces the range of conditions for the evolution of the test-evasive strain. Here, $\alpha_1 = 0.6,\ \alpha_2 = 0.6, \mu = 0.6, d_1 = 0.9$. **(a)** $c = 0, p = \frac{10.9}{10^3}$. **(b)** $c = 0.02$.

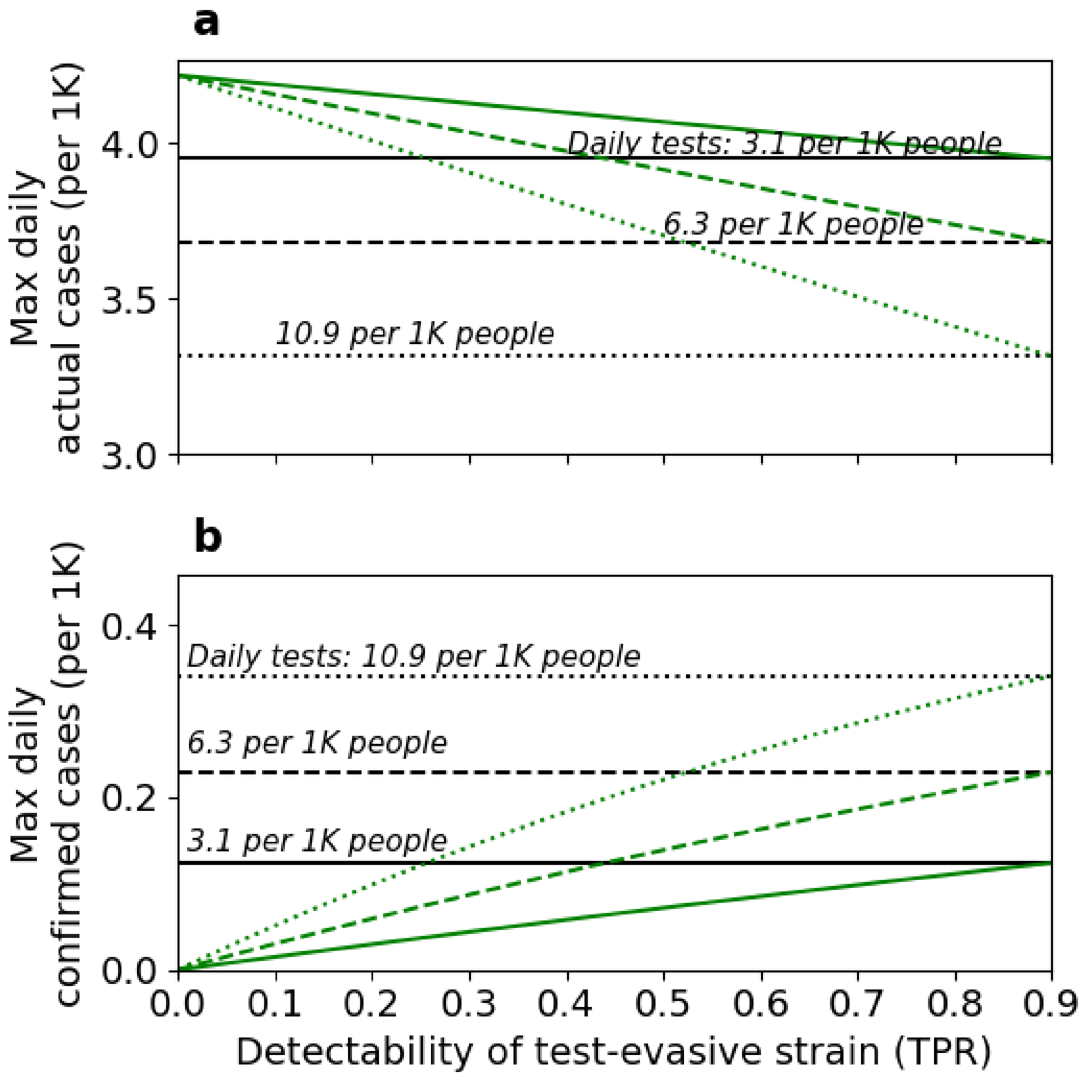


**Figure S4: Effects of decreased detectability without cost of infectiousness on epidemiological metrics.** These results demonstrate the outcomes of two separate epidemic outbreaks: (i) exclusively by a detectable strain (black lines) and (ii) exclusively by a test-evasive strain (green lines), under three testing regimes (solid, dashed, dotted lines). We consider decreased detectability without cost of infectiousness ($c = 0$). These results are similar to Fig. 6 in the main text. However, in this case, the number of actual cases for the test-evasive scenario is always higher than for the detectable strain (compare panel ***a*** here with panel ***a*** in Fig. 6 of the main text). $\beta = 0.42$ (Impact of NPI = 0.65), $\mu = 0.6, \alpha_1 = 0.6, \alpha_2 = 0.6, d_1 = 0.9, c = 0$.

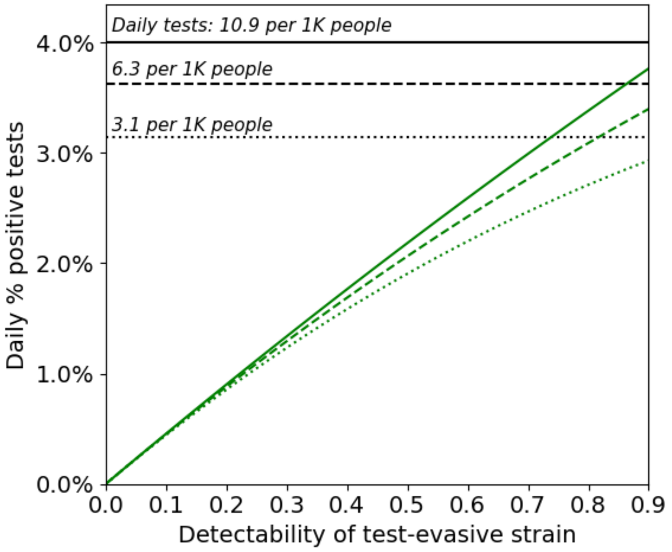


**Figure S5: Effect of decreased detectability on daily percent of positive tests.** These results demonstrate the outcomes of two separate epidemic outbreaks: (i) exclusively by a detectable strain (black lines) and (ii) exclusively by a test-evasive strain (green lines), under three testing regimes (solid, dashed, dotted lines). Daily % of positive tests increases with testing rate, however only mildly when detectability is low (compare solid, dashed, dotted lines). Daily % of positive tests also increases with detectability (green lines). Here, $\beta = 0.42$ (Impact of NPI = 0.65), $\mu = 0.6, \alpha_1 = 0.6, \alpha_2 = 0.6, d_1 = 1, c = 0.01$.
